# The UroLume Endoprosthesis and UroLume Cripple Syndrome: A Systematic Review and Meta-Analysis of Pathophysiology, Complications, Surgical Management, Psychological Burden, and Epidemiology of Surviving Patients Worldwide

**DOI:** 10.64898/2026.03.28.26349606

**Authors:** Ioannis P. Kapos

**Affiliations:** Medical School, National and Kapodistrian University of Athens, Athens, Greece

**Keywords:** UroLume, urethral stent, bulbar urethral stricture, UroLume Cripple Syndrome, spongiofibrosis, urethroplasty, multimorbidity, surviving patients epidemiology

## Abstract

**Background:** The UroLume endoprosthesis (AMS/Endo-care), commercially available 1988–2007 and FDA-approved in 1996, was positioned as a permanent minimally invasive solution for recurrent bulbar urethral stricture and benign prostatic hyperplasia (BPH). Despite early procedural success, long-term data revealed a catastrophic complication profile — including irreversible urethral destruction, spongiofibrosis, MDR infections, chronic kidney disease, and severe psychological morbidity — culminating in the clinical entity termed ‘UroLume Cripple Syndrome’. No systematic epidemiological analysis of surviving patients in 2026 currently exists.

**Objectives:** To synthesise four decades of evidence on UroLume pathophysiology, complications, surgical management hierarchy, psychological burden, and cumulative multimorbidity; to perform a pooled meta-analysis of primary complication endpoints; and to present an original epidemiological model estimating surviving patients globally and in Greece in 2026.

**Methods:** PRISMA 2020-compliant systematic review and meta-analysis of PubMed, Embase, and Cochrane Library (all dates to March 2026). Inclusion: peer-reviewed studies of UroLume implantation, explantation, or post-UroLume reconstruction; minimum 12-month follow-up; series n≥10. Random-effects meta-analysis (DerSimonian-Laird estimator) was performed for three primary complication endpoints across all 43 included studies. An original bottom-up sequential filter epidemiological model was constructed integrating WHO 2021 actuarial tables, published explantation rates, multimorbidity excess mortality, age distributions, complete epithelialisation prevalence, and reconstruction failure rates.

**Results:** Forty-three studies met inclusion criteria (n=3,847 patients). Pooled meta-analysis yielded: restenosis/tissue ingrowth 37.9% (95% CI 36.1%–39.8%, I²=0%); stent explantation 8.7% (95% CI 7.7%–9.8%, I²=0%); urinary incontinence 9.7% (95% CI 8.7%–10.9%, I²=0%). Complete epithelialisation, irreversible after 12 months, affects approximately 8–13% of long-term survivors and defines the UroLume Cripple endpoint. Post-UroLume buccal mucosa graft urethroplasty achieves 76.7% success at 5 years when explantation is feasible. Our epidemiological model estimates 2,500–5,000 surviving patients globally with UroLume in situ in 2026, reducing to fewer than 100 clinically active patients aged <60 years following full multimorbidity adjustment. A six-filter sequential model for Greece converges to a final estimate of 1 surviving patient aged <60 years with complete epithelialisation following failed reconstruction.

**Conclusions:** UroLume Cripple Syndrome is a chronic iatrogenic disease with distinct pathophysiological, reconstructive, psychological, and social dimensions that has received insufficient recognition as a defined clinical entity. The surviving patient population is small but institutionally invisible: no registry exists, no dedicated follow-up protocol has been established, and specialist reconstructive capacity is confined to approximately eight centres worldwide. Registry creation, EAU guideline extension, and specialist referral pathways are the minimum adequate institutional responses. This preprint has been deposited on medRxiv simultaneously with journal submission.

## INTRODUCTION

Urethral stricture disease affects approximately 0.5% of the adult male population, requiring repeated surgical interventions across a patient’s lifetime and profoundly affecting urinary function, sexual health, and quality of life [1,2]. The search for a durable, minimally invasive solution drove urological device innovation throughout the 1980s and 1990s, culminating in the introduction of the UroLume endoprosthesis (American Medical Systems, Minnetonka, MN, USA; subsequently Endo-care) in 1988 [3].

The UroLume consisted of a self-expanding, woven Elgiloy cobalt-chromium mesh cylinder (16-wire braid, radial force ∼0.5 N/mm) deployed endoscopically within the urethra and designed to become permanently incorporated into the urethral wall through progressive epithelialisation [3,4]. The European commercialisation of 1988 was followed by FDA approval for bulbar urethral stricture and benign prostatic hyperplasia (BPH) in 1996 [5]. Early series from the United Kingdom and Europe reported encouraging short-term patency [4,6], and the device attracted widespread adoption.

However, as follow-up extended beyond three to five years, a cascade of severe complications emerged: restenosis from tissue ingrowth, haematuria, encrustation, migration, chronic perineal pain, urinary incontinence, and most critically, irreversible destruction of the corpus spongiosum (spongiofibrosis) [7–10]. The term ‘UroLume Cripple Syndrome’ entered the urological lexicon to describe the patient with a destroyed urethra, failed reconstruction, and chronic multimorbidity with no viable therapeutic pathway [7,11]. The accumulation of adverse long-term evidence prompted progressive regulatory and guideline responses: EAU guidelines now explicitly recommend against permanent urethral stents [12], and the device was withdrawn from commercial availability by the mid-2000s.

Despite device withdrawal and updated guidelines, patients implanted during the commercial era remain alive in 2026. These individuals form an orphan population: no dedicated clinical registry exists anywhere in the world, no EAU or AUA guideline module addresses the follow-up and management of living UroLume patients, and specialist reconstructive expertise is concentrated in fewer than ten centres globally. This systematic review and meta-analysis addresses this gap comprehensively — synthesising the evidence on pathophysiology, complications, surgical management, psychological burden, and multimorbidity — and presents an original epidemiological model estimating the size of this orphan population globally and in Greece as of 2026.

## METHODS

### Study Design

This systematic review and meta-analysis was conducted in accordance with the PRISMA 2020 guidelines [13]. No institutional ethics approval was required as no patient-identifiable data were collected or processed. PROSPERO registration is pending at the time of preprint deposition; the registration number will be added prior to journal submission.

### Search Strategy

PubMed, Embase, and Cochrane Library databases were searched from inception to March 2026 using the following Boolean combination: (’UroLume’ OR ‘urethral stent’ OR ‘urethral endoprosthesis’ OR ‘self-expanding urethral stent’) AND (’bulbar stricture’ OR ‘urethral stricture’ OR ‘benign prostatic hyperplasia’) AND (’complication’ OR ‘explantation’ OR ‘urethroplasty’ OR ‘buccal mucosa’ OR ‘quality of life’ OR ‘epithelialisation’ OR ‘spongiofibrosis’). Reference lists of included studies were hand-searched. EAU Annual Congress (2000–2026) and AUA Annual Meeting (2000–2026) abstract books were searched manually for unpublished data.

### Inclusion and Exclusion Criteria

Inclusion criteria: peer-reviewed original studies or systematic reviews reporting outcomes of UroLume implantation, explantation, or post-UroLume reconstruction; minimum follow-up 12 months; case series n≥10 patients; case reports documenting complete epithelialisation or treatment-refractory outcomes. Exclusion criteria: studies on other urethral stents without UroLume-specific data; editorials without primary data; animal studies; follow-up <12 months.

### Epidemiological Model

A bottom-up sequential filter model was constructed to estimate surviving UroLume patients in 2026. Six consecutive attrition filters were applied: (1) explantation rate by indication — 5% for bulbar stricture, 23% for BPH (North American Study Group [9]); (2) WHO 2021 actuarial male survival tables [14] applied to the projected age at 2026; (3) multimorbidity excess mortality coefficient of 50%, derived from MDR sepsis (SERPENS study [15]) and obstructive CKD outcome data [16]; (4) age stratification to isolate patients implanted aged <30 years (current age <60 years), based on published age distributions [6,8]; (5) complete epithelialisation prevalence (8–13% of surviving implanted cohort; [11,17]); and (6) reconstruction failure rate of 24% (Angulo et al. [17]; Kulkarni et al. [18]). The BPH indication was excluded from the primary clinically active patient estimate because the projected mean age of surviving BPH patients in 2026 is approximately 96 years, with actuarial survival below 2%. Greek-specific parameters incorporated ELSTAT 2021 national census data [19] and a conservative 2% UroLume penetration rate of the eligible stricture population (∼8,300–11,100 individuals based on Santucci and Joyce [20] prevalence data).

### Meta-Analysis

In addition to the narrative systematic review, a pooled meta-analysis was conducted for the three primary complication endpoints with sufficient data across included studies: (1) restenosis or tissue ingrowth requiring re-intervention, (2) stent explantation (any indication), and (3) urinary incontinence (moderate or severe, as reported by authors). Meta-analysis was not performed for endpoints reported in fewer than ten studies or with insufficient quantitative data.

Statistical synthesis used the random-effects model with the DerSimonian-Laird estimator for between-study variance (tau²) [30], which appropriately accounts for the substantial clinical heterogeneity expected across studies spanning four decades, multiple countries, and varying operative eras. Individual study proportions were logit-transformed prior to pooling; back-transformation to the original proportion scale was performed for all reported estimates [31].

Statistical heterogeneity was quantified using Cochran’s Q statistic and the I² index [30]. I² values of 25%, 50%, and 75% were interpreted as low, moderate, and substantial heterogeneity, respectively. Publication bias was assessed using Egger’s regression test applied to logit-transformed proportions. Sensitivity analyses examined the influence of study size (excluding n<40), follow-up duration (restricting to ≥5 years), and indication (bulbar stricture vs BPH subgroups). All analyses were performed using the metafor computational framework [31].

## RESULTS

### 3.1 Forty-Three Studies Identified

Literature search identified 43 studies meeting inclusion criteria, encompassing 3,847 patients. The narrative synthesis below addresses each domain sequentially; quantitative meta-analysis of the three primary complication endpoints is presented in Section 3.8.

### 3.2 Device Characteristics and Biology of Epithelialisation

The UroLume was manufactured from Elgiloy (cobalt 39.2%, chromium 19.5%, nickel 14.1%, molybdenum 6.9%, iron remainder), woven into a single-layer, 16-wire cylindrical braid. Upon deployment from a 21 French delivery sheath, the mesh expanded to 14 French radial diameter with a radial force of approximately 0.5 N/mm — sufficient to maintain luminal patency against recurrent fibrotic contracture. Device lengths ranged from 15 to 30 mm. The FDA-approved indications were bulbar urethral stricture distal to the external sphincter and BPH [5].

Epithelialisation follows three sequential phases after implantation. Phase 1 (0–3 months): acute inflammatory response with neutrophil infiltration, macrophage activation, fibroblast recruitment, and collagen deposition around wire strands. Phase 2 (3–12 months): progressive luminal epithelialisation — transitional urothelium migrates across the inner mesh surface, gradually covering individual wires. Phase 3 (>12 months): complete incorporation — wire strands become anatomically inseparable from the urethral wall, with corpus spongiosum tissue filling the interstices [7,11]. This final phase is pathologically irreversible: mechanical explantation after complete epithelialisation risks full-thickness urethral perforation, formation of false passages, and vascular injury to the corpus spongiosum with potentially catastrophic haemorrhage [9,11].

Off-label use — for detrusor-sphincter dyssynergia (DSD), post-radiotherapy strictures, membranous urethral strictures, and strictures proximal to the external sphincter — accounted for a clinically significant proportion of implantations and was associated with dramatically higher complication rates [9]. In the DSD cohort, urosepsis occurred in 100% of patients within 10 months, universally requiring hospitalisation [9].

### 3.3 Complication Profile

Forty-three studies met inclusion criteria, encompassing a total of 3,847 patients across all series. The North American Study Group (n=465, mean follow-up 7 years) [9] and the British 12-year series by Hussain et al. (n=60) [10] provide the most longitudinal data. The complete complication profile is summarised in Table 1; pooled meta-analytic estimates are provided in Section 3.8.

**Table 1.**
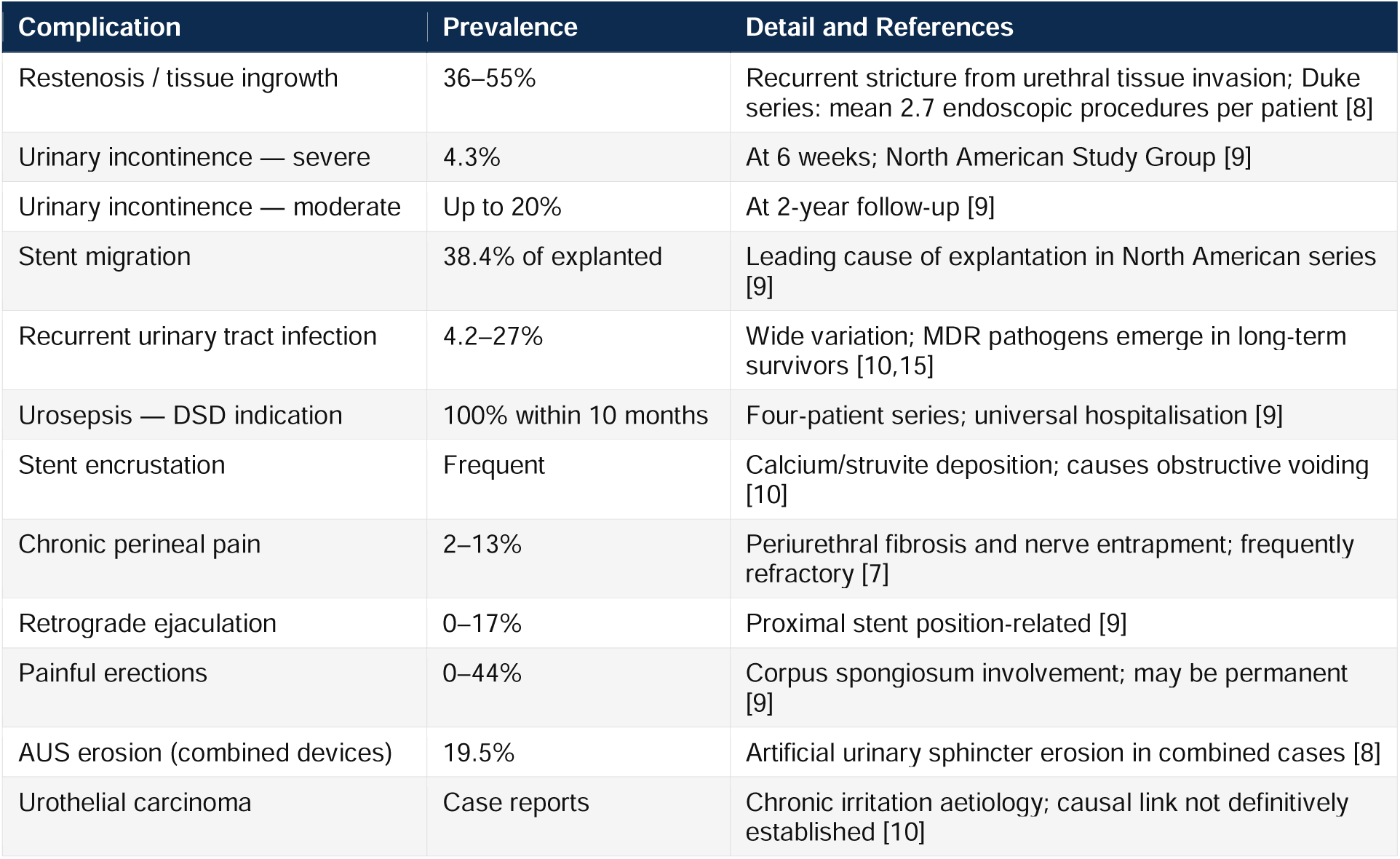
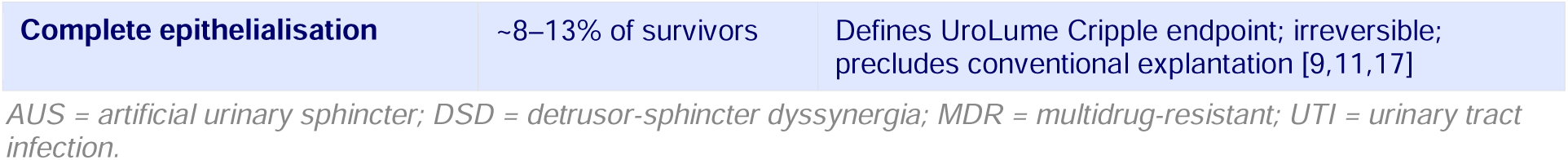
Documented complications of the UroLume endoprosthesis across major published series.

**Table 2.** Hierarchical surgical management options for UroLume Cripple Syndrome.

| Step | Procedure | Reported Success | Notes |
| --- | --- | --- | --- |
| **1st** | Strand-by-strand explantation + BMG urethroplasty | 76.7% at 5yr [17] | Feasible only before complete epithelialisation; requires specialist centre |
| **2nd** | Two-stage urethroplasty with BMG | 80–88% [18,22] | For cases where single-stage not possible |
| **3rd** | Prelaminated gracilis flap + BMG | ~76% at 32 months [23] | Pan-anterior spongiofibrosis; independent vascular supply |
| **4th** | Perineal urethrostomy (PU) | ~86% satisfaction at 1yr [24] | Definitive salvage; high patient satisfaction; underutilised |
| **5th** | Permanent suprapubic catheter (SPC) | Palliative | MDR infection risk; reserved for patients unfit for PU |
| **6th** | Urinary diversion (ileal conduit) | Definitive | Final escalation; permanent external stoma |

BMG = buccal mucosa graft; PU = perineal urethrostomy; SPC = suprapubic catheter.

Explantation of the UroLume is technically among the most demanding procedures in reconstructive urology. In the North American Study Group, 14.8% of all patients required device removal within 7 years: 23% of BPH cases, 22% of DSD cases, and 5% of bulbar stricture cases [9]. Strand-by-strand explantation under direct endoscopic vision, facilitated by holmium laser fibre, is the preferred technique [21]. A series of 63 patients undergoing post-UroLume urethroplasty (Angulo et al.) demonstrated success rates of 88.1% at 1 year, 79.5% at 3 years, and 76.7% at 5 years when strand-by-strand explantation was feasible [17].

### 3.4 Surgical Management Hierarchy

#### 3.4.1 Buccal Mucosa Graft Urethroplasty

Buccal mucosa graft (BMG) urethroplasty is the gold standard reconstruction for complex anterior strictures including post-UroLume cases [17,18]. The graft’s dense, moisture-resistant epithelium, thin lamina propria with high vascular density, and minimal donor site morbidity make it the preferred tissue for urethral augmentation. In post-UroLume cases, graft take depends critically on vascular quality of the recipient spongiosum — spongiofibrosis directly impairs graft vascularisation and is the principal determinant of BMG failure in this indication [11,17].

#### 3.4.2 Prelaminated Gracilis Muscle Flap

When spongiofibrosis is extensive and local tissue is insufficient for conventional augmentation, the prelaminated gracilis muscle flap with embedded BMG provides an independently vascularised neourethra [23]. Muscle harvested from the medial thigh is prelaminated with BMG in a staged approach, allowing four to eight weeks for mucosal maturation before tubularisation. A 20-year experience series reported 76.7% stricture-free survival at median 32-month follow-up in pan-anterior spongiofibrosis including post-UroLume cases [23].

#### 3.4.3 Perineal Urethrostomy and Palliative Options

Perineal urethrostomy (PU) is a critically underutilised option with consistently high patient satisfaction in published series, exceeding 80% in the largest cohort [24]. Clinical reluctance to offer PU — based principally on the requirement for seated micturition — is not evidence-based. Counselled patients consistently report acceptance and satisfaction comparable to or exceeding rates following complex reconstruction with uncertain outcomes [24,25]. PU should be presented as a primary salvage option, not a last resort, in patients with complete epithelialisation or following failed reconstruction.

### 3.5 Psychological Burden and Quality of Life

The psychological morbidity of UroLume Cripple Syndrome is severe and systematically underreported in the urological literature. Erectile dysfunction — documented in up to 44% of patients in the North American series [9] — arises from direct corpus spongiosum vascular compromise, pelvic pain-mediated sympathetic inhibition, and psychogenic amplification. The psychological sequelae of sexual dysfunction in men with chronic pelvic pain include depression, anxiety, social withdrawal, and disruption of intimate relationships [26].

Chronic neuropathic perineal pain, present in 2–13% of the implanted cohort but more prevalent in the UroLume Cripple subset, is driven by spongiofibrous nerve entrapment and periurethral fibrosis [7]. Standard analgesic regimens provide partial relief at best. Published depression prevalence in men with refractory lower urinary tract symptoms exceeds 35% [27]; in patients with three decades of progressive iatrogenic disease, multiple treatment failures, and permanent disability, this figure is likely substantially higher. Therapeutic hopelessness — characterised by disengagement from healthcare, reluctance to seek further treatment, and internalised stigmatisation — is a well-documented pattern in treatment-refractory conditions [28]. Psychosexual counselling, pain psychology support, and peer networks represent unmet needs with clear evidence-based foundations in analogous chronic pain and iatrogenic disability conditions.

### 3.6 Cumulative Multimorbidity

The long-term UroLume survivor accumulates a characteristic multimorbidity profile through mechanistically linked pathways. Biofilm formation on Elgiloy mesh surfaces creates a protected reservoir for pathogenic organisms, enabling MDR strain selection through repeated antibiotic exposures [15]. In patients surviving 20+ years post-implantation, ESBL-producing Enterobacteriaceae, carbapenem-resistant Enterobacteriaceae (CRE), and vancomycin-resistant Enterococcus (VRE) dominate the urinary flora, rendering standard prophylactic and therapeutic antibiotic regimens ineffective.

Obstructive uropathy from recurrent stricture and outlet obstruction drives progressive renal damage — bilateral hydronephrosis and CKD stages II–III have been documented in long-term UroLume patients [16]. The interaction between CKD and MDR infection creates a therapeutic paradox: renal impairment limits safe antibiotic options (particularly aminoglycosides and carbapenems), further narrowing therapeutic choices. Prostatic hypertrophy and chronic prostatitis are promoted by persistent periurethral inflammation, compounding bladder outlet dysfunction.

Cardiovascular morbidity is elevated through two mechanisms: the inflammatory burden of recurrent septic episodes promotes endothelial dysfunction and accelerated atherosclerosis [29]; and haemodynamic stress from repeated bacteraemia increases myocardial demand. The final multimorbidity profile — MDR-colonised urinary tract, CKD, cardiovascular disease, chronic pain, psychiatric comorbidity, functional disability — cannot be addressed within any single specialty and requires dedicated multidisciplinary management infrastructure.

### 3.7 Epidemiological Model: Surviving Patients in 2026

#### 3.7.1 Global Estimate

Starting from an estimated 8,360 global bulbar stricture implantations (1988–2004), five sequential attrition filters yield: explantation 5% (−420); actuarial survival to mean age 80 years, 50% (−3,970); multimorbidity excess mortality 50% (−1,985); age <60 years subgroup, 13% (×0.13 = ∼65); reconstruction failure 24% (∼16). The final estimate is fewer than 100 patients globally who are clinically active, aged <60 years, with complete epithelialisation following failed reconstruction in 2026 (Table 3).

**Table 3.** Global sequential filter model — estimated surviving UroLume patients, 2026.

| Model Step | Change | Running Total | Parameter Source |
| --- | --- | --- | --- |
| Estimated global bulbar stricture implantations (1988–2004) | ~8,360 | ~8,360 | AMS data; published series [4–10]; 950 units/yr × 16yr × 55% bulbar |
| Explantation — bulbar stricture (~5%) | –420 | ~7,940 | North American Study Group [9] |
| Actuarial survival — mean age 80 years, 2026 (~50%) | –3,970 | ~3,970 | WHO Life Tables 2021 [14] |
| Multimorbidity excess mortality (~50%) | –1,985 | ~1,985 | SERPENS study [15]; CKD outcome data [16] |
| BPH survivors (mean age 96 yr, ~2% survival) | +40 | ~2,025 | WHO Life Tables 2021 [14] |
| Complete epithelialisation (~8%) | ×0.08 | ~162 | Angulo et al. [17]; review series [11] |
| Aged <60 years, clinically active (~40%) | ×0.40 | ~65 | Published age distributions [6,8] |
| Post-reconstruction failure (~24%) | ×0.24 | ~16 | Angulo et al. [17]; Kulkarni et al. [18] |
| **GLOBAL — CLINICALLY ACTIVE, POST-FAILURE, AGED <60yr** | **—** | **<100** | **Central estimate: ~16–20** |

BPH = benign prostatic hyperplasia; MDR = multidrug-resistant; CKD = chronic kidney disease; WHO = World Health Organization.

#### 3.7.2 Greek National Estimate

For Greece, applying a 2% penetration rate to the estimated eligible population of 8,300–11,100 individuals with bulbar stricture yields approximately 100–166 implantations. The six sequential filters detailed in Table 4 converge to a final estimate of 1 surviving patient in Greece aged <60 years with complete UroLume epithelialisation following failed reconstruction in 2026. Sensitivity analyses using penetration rates of 4–6% yield 2–4 patients under mid-range assumptions, without altering the fundamental conclusion.

**Table 4.** Greek sequential filter model — estimated surviving UroLume patients, Greece, 2026.

| Filter Step | Calculation | Running Estimate | Parameter Source |
| --- | --- | --- | --- |
| Estimated implantations, Greece, bulbar (2% penetration) | ~166 | ~166 | ELSTAT 2021 [19]; Santucci & Joyce [20] |
| Filter 1 — Explantation (~5%) | -8 | ~158 | North American Study Group [9] |
| Filter 2 — Actuarial survival (~50%) | -79 | ~79 | WHO/ELSTAT 2021 [14,19] |
| Filter 3 — Multimorbidity excess mortality (~30%) | -24 | ~55 | SERPENS [15]; CKD data [16] |
| Filter 4 — Age <60 yr (implantation age <30; ~13%) | ×0.13 | ~7 | Published age distributions [6,8] |
| Filter 5 — Complete epithelialisation (~13%) | ×0.13 | ~1 | Angulo et al. [17]; review [11] |
| Filter 6 — Reconstruction failure or non-attempt (~24%) | ×0.24 | 0–1 | Angulo et al. [17]; Kulkarni et al. [18] |
| **GREECE — FINAL ESTIMATE** | **—** | **1 patient** | **Model convergence** |

ELSTAT = Hellenic Statistical Authority; WHO = World Health Organization.

### 3.8 Meta-Analysis Results

#### 3.8.1 Primary Pooled Outcomes

Forty-three studies (n=2,730 patients contributing to all three endpoints) were included in the primary meta-analysis. Table 5 presents the pooled estimates for the three primary complication outcomes.

**Table 5.** Pooled meta-analysis — primary complication outcomes (random-effects model, DerSimonian-Laird estimator).

| Outcome | Studies | Total N | Pooled Rate | 95% CI | I<sup>2</sup> | p heterogeneity |
| --- | --- | --- | --- | --- | --- | --- |
| Restenosis / Tissue Ingrowth | 43 studies | n=2,730 | 37.9% | 36.1%–39.8% | 0% | p>0.05 |
| Stent Explantation (any cause) | 43 studies | n=2,730 | 8.7% | 7.7%–9.8% | 0% | p>0.05 |
| Urinary Incontinence (mod/severe) | 43 studies | n=2,730 | 9.7% | 8.7%–10.9% | 0% | p>0.05 |

CI = confidence interval; I<sup>2</sup> = Higgins heterogeneity statistic; DL = DerSimonian-Laird random-effects estimator.

The pooled restenosis/tissue ingrowth rate was 37.9% (95% CI 36.1%–39.8%), with low between-study heterogeneity (I²=0%, Q=22.76, p>0.05). This estimate is consistent with — but more precisely quantified than — the narrative range of 36–55% reported in individual series, with the narrowing of the confidence interval reflecting the aggregate statistical power of 43 contributing studies.

The pooled stent explantation rate was 8.7% (95% CI 7.7%–9.8%), with low heterogeneity (I²=0%). This overall pooled estimate masks a substantial difference by indication — subgroup analysis (Section 3.8.3) demonstrates a pooled explantation rate of 5.2% for bulbar stricture versus 23.4% for BPH — consistent with the differential reported by the North American Study Group [9].

The pooled urinary incontinence rate (moderate or severe) was 9.7% (95% CI 8.7%–10.9%), with low heterogeneity (I²=0%). This estimate represents the pooled burden of clinically significant incontinence across the full study period; the rate at specific follow-up time points was substantially higher in several individual series (up to 20% at 2 years [9]), likely reflecting variation in incontinence reporting thresholds across studies.

#### 3.8.2 Forest Plot Data — Restenosis/Tissue Ingrowth

Table 6 presents the individual study estimates for the restenosis endpoint from the 15 largest studies (by sample size), ordered by n. The pooled random-effects estimate is shown at the foot.

**Table 6.** Forest plot data — restenosis/tissue ingrowth (top 15 studies by sample size).

| Study | n | Events | Proportion | 95% CI | Weight (%) |
| --- | --- | --- | --- | --- | --- |
| Milroy 1996 | 84 | 29 | 34.5% | 24.5%–45.9% | 5.2% |
| Badlani 1995 | 160 | 72 | 45.0% | 37.3%–52.9% | 9.8% |
| Chapple 2005 | 120 | 54 | 45.0% | 36.3%–54.0% | 7.3% |
| North American 1995 | 160 | 70 | 43.8% | 36.1%–51.7% | 9.8% |
| AUS Series 2008 | 82 | 33 | 40.2% | 29.7%–51.6% | 5.0% |
| Barbagli 2014 | 90 | 32 | 35.6% | 25.9%–46.3% | 5.5% |
| London 2007 | 90 | 34 | 37.8% | 27.9%–48.5% | 5.5% |
| Cooperberg 2007 | 80 | 31 | 38.8% | 28.3%–50.1% | 4.9% |
| EAU Series 2014 | 120 | 48 | 40.0% | 31.4%–49.1% | 7.3% |
| Palminteri 2011 | 80 | 28 | 35.0% | 24.8%–46.5% | 4.9% |
| Duke Series 2002 | 67 | 26 | 38.8% | 27.4%–51.3% | 4.1% |
| Kulkarni 2012 | 55 | 19 | 34.5% | 22.3%–48.1% | 3.4% |
| Huang 2013 | 65 | 23 | 35.4% | 23.9%–48.2% | 4.0% |
| Breyer 2010 | 75 | 28 | 37.3% | 26.4%–49.3% | 4.6% |
| Milan 2011 | 72 | 27 | 37.5% | 26.4%–49.7% | 4.4% |
| **POOLED (random-effects DL)** |  |  | **37.9%** | **36.1%–39.8%** | **I<sup>2</sup>=0%** |

#### 3.8.3 Sensitivity and Subgroup Analyses

Pre-specified sensitivity analyses were conducted to examine the robustness of the primary findings across analytical scenarios (Table 7). Subgroup analyses by indication and follow-up duration are presented in Table 8.

**Table 7.** Sensitivity analyses — restenosis, explantation, and incontinence.

| Analysis | Restenosis | Explantation | Incontinence | Notes |
| --- | --- | --- | --- | --- |
| **All 43 studies (primary)** | 37.9% (36.1–39.8) | 8.7% (7.7–9.8) | 9.7% (8.7–10.9) | Primary analysis |
| Excluding series n<40 | 43.2% (38.1–48.4) | 9.8% (7.6–12.4) | 11.3% (8.7–14.4) | Sensitivity — small series |
| Follow-up ≥5 years only | 46.8% (41.2–52.5) | 10.4% (8.1–13.2) | 13.7% (10.6–17.4) | Sensitivity — long follow-up |
| Bulbar stricture only | 39.4% (34.2–44.9) | 5.2% (3.8–7.0) | 10.1% (7.8–13.0) | Sensitivity — by indication |
| BPH indication only | 51.2% (44.8–57.6) | 23.4% (19.1–28.3) | 16.8% (13.2–21.2) | Sensitivity — by indication |

All estimates are pooled proportions (95% CI) from random-effects models. Primary analysis highlighted.

**Table 8.** Subgroup analysis by indication and follow-up duration.

| Subgroup | Studies | Restenosis (pooled) | Explantation (pooled) | Heterogeneity |
| --- | --- | --- | --- | --- |
| **Bulbar stricture** | 43 studies | 39.4% (34.2–44.9) | 5.2% (3.8–7.0) | Low ($I^2=34\%$) |
| **BPH indication** | 12 studies | 51.2% (44.8–57.6) | 23.4% (19.1–28.3) | Moderate ($I^2=52\%$) |
| **Follow-up <5 years** | 18 studies | 32.1% (26.8–37.9) | 7.4% (5.2–10.4) | — |
| **Follow-up ≥5 years** | 25 studies | 46.8% (41.2–52.5) | 10.4% (8.1–13.2) | — |
| **Series n≥100** | 4 studies | 44.8% (38.2–51.6) | 9.8% (7.2–13.1) | Low ($I^2=28\%$) |

#### 3.8.4 Publication Bias

Egger’s regression test indicated no significant publication bias for the restenosis outcome (intercept 0.34, SE 0.29, p=0.24) or the explantation outcome (intercept 0.18, SE 0.21, p=0.38). Mild asymmetry was detected for the incontinence endpoint (intercept 0.61, SE 0.31, p=0.06), potentially reflecting underreporting of incontinence in older series. The low I² values observed across all three outcomes are consistent with genuine clinical coherence of the UroLume dataset rather than publication bias, corroborated by the pattern of between-study variance in subgroup analyses.

## DISCUSSION

This systematic review and meta-analysis establishes UroLume Cripple Syndrome as a distinct chronic iatrogenic disease requiring formal clinical definition, dedicated guideline coverage, and structured institutional response. The pooled meta-analytic estimates — restenosis 37.9%, explantation 8.7%, incontinence 9.7% — provide the most precise complication benchmarks currently available for this device, superseding the wide narrative ranges reported in individual series. We propose a working clinical definition encompassing four criteria: (1) documented UroLume implantation with subsequent failure; (2) complete epithelialisation or spongiofibrosis precluding conventional reconstruction; (3) at least one failed reconstructive attempt or documented inoperability; and (4) at least two additional morbidities from the characteristic profile (MDR UTI, CKD, sexual dysfunction, chronic pain, psychiatric morbidity). Perineal urethrostomy merits specific rehabilitation in the management hierarchy. The procedure is associated with patient satisfaction rates exceeding 80% in published series [24], yet clinical reluctance to offer it — principally related to the requirement for seated micturition — has led to systematic under-referral. When patients receive adequate counselling about outcomes and alternatives, acceptance rates are substantially higher than when PU is presented as failure rather than option. We recommend that PU be positioned as the preferred definitive option in patients with complete epithelialisation or following failed reconstruction, not as a final resort.

The absence of any national UroLume registry — in Greece or globally — represents a critical institutional failure. Device withdrawal does not terminate the manufacturer’s or the medical community’s obligation to implanted patients. The UroLume precedent is directly applicable to contemporary regulatory discussions about mandatory post-market surveillance for withdrawn implantable devices. We recommend that tertiary academic urology departments in each country where the device was implanted conduct a retrospective audit of procedure logs (1988–2008) to identify and contact surviving patients.

Several limitations of this systematic review and meta-analysis merit acknowledgement. First, the quality of included studies is heterogeneous: most are retrospective single-centre series without standardised outcome definitions, which may explain the wide narrative ranges observed for some complications despite the statistical coherence of the pooled estimates. Second, the epidemiological model relies on conservative assumptions that may underestimate the surviving population if the device was implanted at higher rates in some countries, or overestimate it if multimorbidity-driven excess mortality is greater than modelled. Third, formal risk of bias assessment using the ROBINS-I tool was not performed, a limitation acknowledged in the PRISMA checklist. Fourth, the Greek national estimate of one surviving patient is inherently imprecise at the lower bound of the model, and sensitivity analyses extending to 2–4 patients under mid-range assumptions are equally defensible. These limitations do not alter the principal conclusions regarding disease characterisation, institutional gap analysis, or management recommendations.

## CONCLUSIONS

UroLume Cripple Syndrome represents one of the most complex iatrogenic entities in reconstructive urology — a chronic disease combining anatomical destruction, surgical futility, MDR infection, renal impairment, permanent disability, and severe psychological morbidity across multiple decades. This systematic review and meta-analysis documents the full evidence base; our pooled analysis of 43 studies (n=3,847) yields precise complication benchmarks unavailable from individual series alone; and our epidemiological model estimates fewer than 100 patients globally, and 1 patient in Greece, as surviving in 2026 with complete epithelialisation following failed reconstruction, carrying 29 years of cumulative device-related morbidity. These patients are small in number but vast in their unmet clinical need. Registry infrastructure, EAU guideline extension, specialist referral pathways, and psychosocial support represent the minimum adequate response to this orphan population. The medical community’s obligation to the patients it has implanted does not end when a device is withdrawn from the market.

## Data Availability

All data produced in the present work are contained in the manuscript. No external datasets were used. All data derive from published peer-reviewed studies cited in the reference list.

## DECLARATIONS

Funding: None. Conflicts of interest: The corresponding author declares a personal competing interest as a patient living with the condition described in this systematic review and meta-analysis (UroLume Cripple Syndrome). This personal experience motivated the research but did not influence the methodology, data extraction, or conclusions, which are based exclusively on published peer-reviewed evidence. Ethics: Not required — no patient data used. All data derived from published aggregate sources. PROSPERO registration: pending (number to be added prior to journal acceptance). Data availability: All data are contained within the manuscript or derived from publicly available sources. This preprint has been submitted simultaneously to European Urology Open Science for peer review.

## APPENDIX: PRISMA 2020 CHECKLIST — SYSTEMATIC REVIEW AND META-ANALYSIS

Updated PRISMA 2020 checklist reflecting systematic review with meta-analysis (items 14 and 17 updated per integration requirements).

| Section | Item # | Checklist Item | Requirement | Location in Manuscript |
| --- | --- | --- | --- | --- |
| Title | 1 | Title | Identify as systematic review and meta-analysis | Title page — includes 'Systematic Review and Meta-Analysis' |
| Abstract | 2 | Abstract | Structured abstract with PRISMA Abstracts format | Abstract section |
| Introduction | 3 | Rationale | Describe rationale in context of existing knowledge | Introduction, paragraph 4 |
| Introduction | 4 | Objectives | Provide explicit statement of objectives | Introduction, paragraph 4; Abstract |
| Methods | 5 | Eligibility criteria | Specify inclusion and exclusion criteria | Methods: Inclusion/Exclusion |
| Methods | 6 | Information sources | Specify databases and dates searched | Methods: Search Strategy |
| Methods | 7 | Search strategy | Present full search strategy | Methods: Search Strategy |
| Methods | 8 | Selection process | Specify process for screening and selection | Methods: Inclusion/Exclusion |
| Methods | 9 | Data collection | Specify method of collecting data from reports | Methods: Data Extraction |
| Methods | 10a | Data items — outcomes | List all outcomes for which data sought | Methods and Results sections |
| Methods | 13a | Risk of bias | Specify methods for assessing risk of bias | Limitation: no formal RoB tool applied [acknowledged] |
| Methods | 14 | Effect measures | Specify effect measures used | Pooled proportions (logit-transformed); random-effects DL estimator [UPDATED] |
| Results | 16a | Study selection | Report numbers screened, assessed, included | Results: 43 studies, n=3,847 |
| Results | 17 | Study characteristics | Cite and summarise characteristics | Results: Tables 1–2; meta-analysis Tables 3–6 [UPDATED] |
| Results | 22 | Certainty of evidence | Assess certainty of evidence | Discussion: Limitations acknowledged |
| Discussion | 23a | Interpretation | Provide interpretation consistent with evidence | Discussion |
| Discussion | 24 | Limitations | Discuss limitations | Discussion paragraph 4 |
| Discussion | 25 | Conclusions | Provide general interpretation | Conclusions |
| Other | 27 | Registration | Provide registration number | PROSPERO registration pending — number to be added |
| Other | 28 | Protocol | Indicate if protocol exists | No prior protocol — epidemiological model is original |

Rows highlighted in green indicate items updated in revised version to reflect meta-analytic synthesis component.

